# Development and external validation of the DOAT and DOATS scores: simple decision support tools to identify disease progression among nonelderly patients with mild/moderate COVID-19

**DOI:** 10.1101/2021.12.13.21267698

**Authors:** Yoko Shibata, Hiroyuki Minemura, Yasuhito Suzuki, Takefumi Nikaido, Yoshinori Tanino, Atsuro Fukuhara, Ryuzo Kanno, Hiroyuki Saito, Shuzo Suzuki, Taeko Ishii, Yayoi Inokoshi, Eiichiro Sando, Hirofumi Sakuma, Tatsuho Kobayashi, Hiroaki Kume, Masahiro Kamimoto, Hideko Aoki, Akira Takama, Takamichi Kamiyama, Masaru Nakayama, Kiyoshi Saito, Koichi Tanigawa, Masahiko Sato, Toshiyuki Kanbe, Norio Kanzaki, Teruhisa Azuma, Keiji Sakamoto, Yuichi Nakamura, Hiroshi Otani, Mitsuru Waragai, Shinsaku Maeda, Tokiya Ishida, Keishi Sugino, Minoru Inage, Noriyuki Hirama, Kodai Furuyama, Shigeyuki Fukushima, Hiroshi Saito, Jun-ichi Machiya, Hiroyoshi Machida, Koya Abe, Katsuyoshi Iwabuchi, Yuji Katagiri, Yasuko Aida, Yuki Abe, Takahito Ota, Yuki Ishizawa, Yasuhiko Tsukada, Ryuki Yamada, Riko Sato, Takumi Omuna, Hikaru Tomita, Mikako Saito, Natsumi Watanabe, Mami Rikimaru, Takaya Kawamata, Takashi Umeda, Julia Morimoto, Ryuichi Togawa, Yuki Sato, Junpei Saito, Kenya Kanazawa, Kenji Omae, Kurita Noriaki, Ken Iseki

## Abstract

**BACKGROUND:** Due to the dissemination of vaccination against severe acute respiratory syndrome coronavirus 2 in the elderly, the virus-susceptible subjects have shifted to unvaccinated non-elderlies. The risk factors of COVID-19 deterioration in non-elderly patients without respiratory failure have not yet been determined. This study was aimed to create simple predicting method to identify such patients who have high risk for exacerbation.

**METHODS:** We analyzed the data of 1,675 patients aged under 65 years who were admitted to hospitals with mild-to-moderate COVID-19. For validation, 324 similar patients were enrolled. Disease progression was defined as administration of medication, oxygen inhalation and mechanical ventilator starting one day or longer after admission.

**RESULTS:** The patients who exacerbated tended to be older, male, had histories of smoking, and had high body temperatures, lower oxygen saturation, and comorbidities such as diabetes/obesity and hypertension. Stepwise logistic regression analyses revealed that comorbidities of <u>d</u>iabetes/<u>o</u>besity, <u>a</u>ge ≥ 40 years, body temperature ≥ 38°C, and oxygen saturation < 96% (DOATS) were independent risk factors of worsening COVID-19. As a result two predictive scores were created: DOATS score, which includes all the above risk factors; and DOAT score, which includes all factors except for oxygen saturation. In the original cohort, the areas under the receiver operating characteristic curve of the DOATS and DOAT scores were 0.789 and 0.771, respectively. In the validation, the areas were 0.702 and 0.722, respectively.

**CONCLUSION:** We established two simple prediction scores that can quickly evaluate the risk of progression of COVID-19 in non-elderly, mild/moderate patients.

**Summary:** The risk stratification models using independent risks, namely comorbidity of <u>d</u>iabetes or <u>o</u>besity, <u>a</u>ge ≥ 40 years, high body temperature ≥ 38□, and oxygen saturation < 96%, DOATS and DOAT scores, predicted worsening COVID-19 in patients with mild-to-moderate cases.

---

Due to the global spread of severe acute respiratory syndrome coronavirus 2 (SARS-CoV-2), the coronavirus disease 2019 (COVID-19) pandemic remains a serious problem all over the world. Approximately 5% of COVID-19 patients develop respiratory failure, and 2% die despite undergoing intensive treatment[1]. Even in Japan, many patients could not be admitted to hospital due to the lack of available beds during the height of pandemic in urban areas such as Tokyo, Kobe and Osaka. Therefore, it is important to predict, using simple measures, which patients will become severely ill among those who do not have respiratory failure in the early phase of infection. Until now, some observational studies revealed the risk factors for severe COVID-19. Old age, male sex, smoking habit, and comorbidities such as diabetes, chronic renal disease, malignancies, and chronic respiratory diseases such as chronic pulmonary disease or interstitial lung diseases have been listed as risk factors for severe COVID-19[2-7]. In addition, some biomarkers have been reported to be associated with disease severity[3, 7-11]. Using these risk factors, several investigators reported risk stratification methods[8-10, 12-15].

From February 2021, vaccine against SARS-CoV-2 began to be disseminated in Japan. By the end of July 2021, 30% of residents in Japan were fully vaccinated. Most of them were elderlies and health-care workers. In accordance with the dissemination of the vaccine, risk stratification using risk factors reported may no longer be available. For example, cases in older adults who have been vaccinated are reported to be less severe than cases before the vaccinations were started. Instead, the number of middle-aged individuals who have severe COVID-19 have been increasing, because they haven’t had a chance to receive it yet. Therefore, the subjects who are in most need of risk stratification should be refocused to the young/middle-aged generations, rather than the elderly.

In Fukushima, Japan, by the end of May 2021, approximately 4,500 residents tested positive for SARS-CoV-2 via PCR examination. Twenty-six medical institutes in Fukushima Prefecture began to gather the medical information of all COVID-19 patients who had been admitted to the hospitals, starting from March 2020, and 66% of the COVID-19 patients in Fukushima (3,008 patients) were enrolled by the end of May 2021. The data of not only severely ill patients, but also those of many patients with mild-to-moderate cases are also available in this database. Until now, few investigations have been reported about the risk factors among the general population aged < 65 years who have not been vaccinated for SARS-CoV-2[16]. The aims of the present study were to identify the risk factors for aggravation of COVID-19 among the non-elderly and mild/moderate patients who have not received the vaccine, as well as to develop and externally validate clinical prediction rules in order to identify worsening cases. In particular, we analyzed the factors associated with exacerbation which were identified based on not only death and the initiation of mechanical ventilation, but also the start of medications such as remdesivir or dexamethasone, and the start of oxygen inhalation during hospitalization among COVID-19 patients aged < 65 years who did not have respiratory failure on admission.

## Methods

### Setting and study population

This multicenter retrospective, cohort study used the data of all consecutive patients with COVID-19 admitted to the hospitals that participated in a web conference against COVID-19 held weekly by the Department of Pulmonary Medicine, Fukushima Medical University, between March 31, 2020 and May 20, 2021. Of the 43 COVID-19 hospitals in Fukushima, 26 facilities that engaged in medical care during the acute phase of the disease participated in this conference. Of 4,500 patients with COVID-19 in Fukushima, 3,008 were enrolled in this study. For the external validation study, three hospitals in Yamagata treating COVID-19 patients between March 31, 2020 and May 20, 2021 gave the data of 324 mild and moderate patients aged < 65 years. A diagnosis of COVID-19 was made when the results of a PCR test using a nasopharyngeal swab or saliva were positive. The subjects’ data, such as clinical characteristics including comorbidities, results of examination, medical course, medications, and outcomes, were obtained from the medical records of each hospital. COVID-19 disease severity was classified according to the definition of the Japanese Ministry of Health, Labor and Welfare as follows: mild, subjects without pneumonia and respiratory failure; moderate-1, subjects with pneumonia but without respiratory failure; moderate-2, subjects with pneumonia and respiratory failure (oxygen saturation < 94% on room air) that does not require mechanical ventilation or extracorporeal membrane oxygenation (ECMO); severe, subjects with pneumonia and respiratory failure that required mechanical ventilation and ECMO[17, 18].

Among the 3,008 enrolled patients, 1,675 had mild and moderate-1 cases, and were aged < 65 years. These were the eligible subjects in this study (Figure 1).

**Figure 1.**
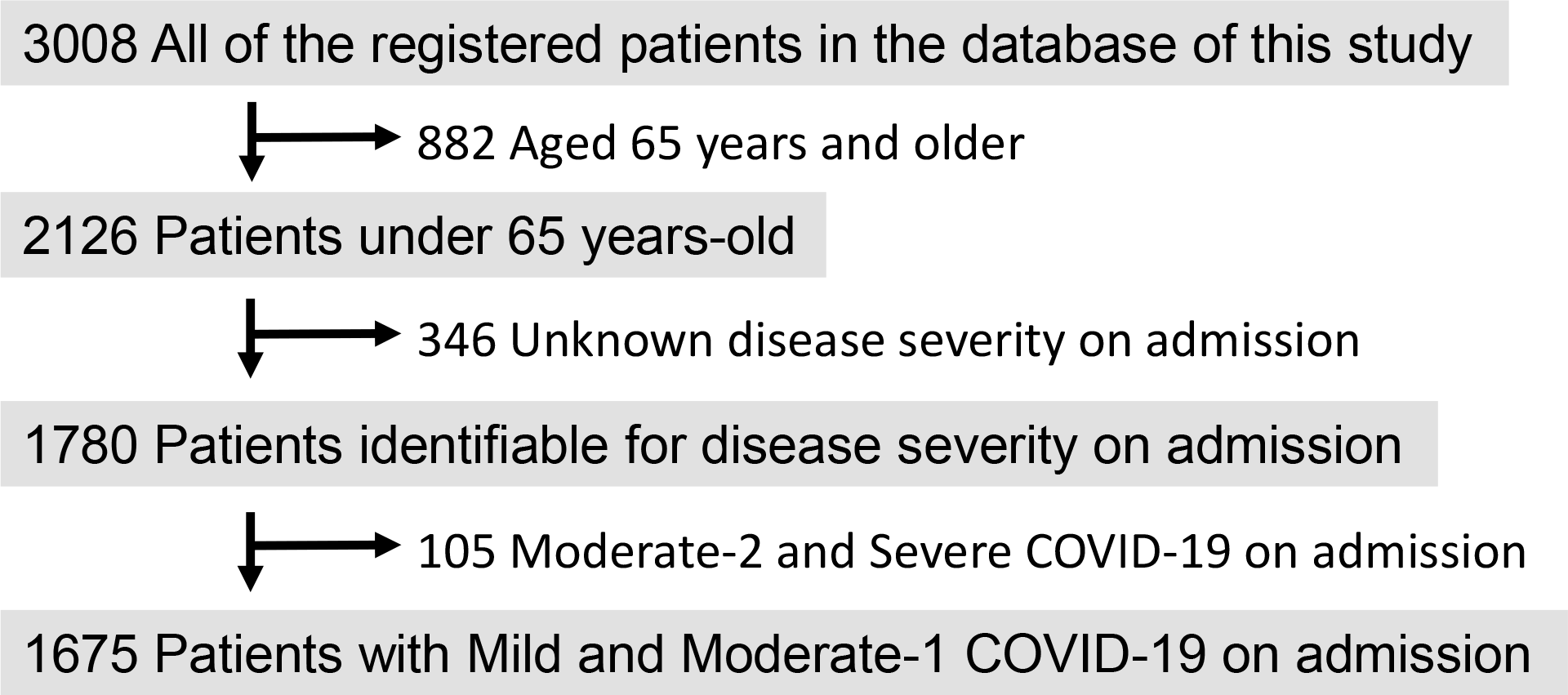
Flowchart of patients’ recruitment in this study. Among all of the registered inpatients with COVID-19 in Fukushima, 1,675 were selected for the present study. Disease severity was classified into mild, moderate-1, moderate-2 and severe in accordance with the definition of Japanese Ministry of Health, Labor and Welfare, as described in the *Methods* section of this manuscript.

### Outcomes

The outcomes were: (1) any disease deterioration, defined as the initiation of medication for COVID-19 (dexamethasone, methylprednisolone, tocilizumab, baricitinib, or remdesivir) or respiratory therapy (use of inhalation oxygen); or (2) use of a ventilator or the need for ECMO after the first day of hospitalization.

### Candidate predictors

Age, sex, body temperature, history of smoking, comorbidities (prevalence 2% and higher), pregnancy, and blood oxygen saturation measured by pulse oximeter were evaluated as candidates for the risk factors for deterioration of the disease. The collection of information was left to the doctors at each participating institution.

### Statistical analyses

Continuous variables are presented as medians with interquartile ranges, and categorical variables are presented as number of patients with percentage. Patients whose conditions exacerbated a day or later after admission were assigned to the Worsened group and the other to the Stable group. Between-group comparisons were performed using the Mann-Whitney U test for the continuous variables and the chi square test for the categorical values. The continuous variables, age and oxygen saturation measured by saturation monitor, were turned into categorical variables using the cut-off described below. The cut-off age was set as 40 years old, because that age is considered to be the boundary between young adults and middle-aged people. The cut-off of oxygen saturation was set as 96%[19]. Among the comorbidities, those with a prevalence of 2% or more were applied to the analyses. Because diabetes and obesity share common clinical features and immune responses to SARS-CoV-2[20], and both have been reported as high risk factors for COVID-19 mortality[21], they fit into the same category in order to simplify the prediction model. Other statistical methods are described in the Supplementary Materials.

### Ethics statement

The need for informed consent was waived because the study is retrospective. This study was approved by the Ethical Committee of Fukushima Medical University (approval number 2020-118, approved on August 3, 2020, updated September 01, 2021).

## Results

Table 1 shows the characteristics of the mild/moderate-1 COVID-19 patients aged under 65 years old in the current study. One hundred forty-four patients (8.6%) experienced worsening of the disease a day after admission or later. These patients were older and more male, and had histories of cigarette smoking, higher body temperature, lower oxygen saturation, and comorbidities such as diabetes/obesity, and hypertension. The rates of pregnancy, as well as comorbidities such as chronic respiratory diseases, malignancies, dyslipidemia, and cardiac diseases, were not significantly different between the Worsened and Stable groups.

**Table 1.** Differences of characteristics between the Stable and Worsened groups in the original cohort of the study and external validation cohort.

|  | Original cohort |  |  | External validation cohort |  |  |
| --- | --- | --- | --- | --- | --- | --- |
|  | All<br>n = 1675 | Stable group<br>n = 1531 | Worsened group<br>n = 144 | All<br>n = 324 | Stable group<br>n = 283 | Worsened group<br>n = 41 |
| Age, year | 41 (25–53) | 39 (24–52) | 51 (41.25–59.75)*** | 42 (29.3–52) | 41 (27–51) | 51 (39.5–60.5)### |
| Male | 919 (54.8%) | 825 (53.9%) | 94 (65.3%)*** | 181 (55.9%) | 158 (55.8%) | 23 (56.1%)<sup>a</sup> |
| Mild / Moderate-1 | 947 / 728 | 900 / 631 | 47 / 97 *** | 260 / 64 | 240 / 43 | 20 / 21 ### |
| History of cigarette smoking | 550 (38.0%) | 479 (36.56%) | 71 (52.59%)*** | — | — | — |
| Temperature ≥37°C | 647 (40.4%) | 549 (37.6%) | 98 (68.5%)*** | 230 (71.0%) | 195 (68.9%) | 35 (85.4%)<sup>#</sup> |
| Temperature ≥38°C | 291 (17.8%) | 219 (14.7%) | 72 (50.7%)*** | 96 (29.6%) | 73 (25.8%) | 23 (56.1%)### |
| Diabetes or Obesity | 175 (11.2%) | 132 (9.26%) | 43 (30.5%)*** | 53 (16.6%) | 41 (14.7%) | 12 (29.3%)<sup>#</sup> |
| Hypertension | 197 (12.6%) | 155 (10.87%) | 42 (29.8%)*** | — | — | — |
| Chronic respiratory diseases | 75 (4.8%) | 67 (4.7%) | 8 (5.7%) | — | — | — |
| Malignancies | 37 (2.4%) | 31 (2.2%) | 6 (4.3%) | — | — | — |
| Dyslipidemia | 80 (5.1%) | 68 (4.74%) | 12 (8.51%) | — | — | — |
| Cardiac diseases | 40 (2.5%) | 33 (2.30%) | 7 (4.96%) | — | — | — |
| Pregnancy | 13 (0.8%) | 13 (0.8%) | 0 (0%) | — | — | — |
| MV and/or ECMO | 7 (0.4%) | 0 (0%) | 7 (0.4%) | — | — | — |
| Deceased due to COVID-19 | 1 (0.06%) | 0 (0%) | 1 (0.06%) | — | — | — |
| SpO<sub>2</sub>, % | 97 (97–98) | 98 (97–98) | 97 (96–97.75)*** | 97 (96–98) | 97 (97–98) | 97 (96–98)<sup>b</sup> |

\*\*\*: $P < 0.0001$ vs Stable group (original cohort), ###: $P < 0.0001$ vs Stable group (validation cohort), #: $P < 0.05$ vs Stable group (validation cohort), <sup>a</sup>: $P = 0.974$ vs Stable group (validation cohort), <sup>b</sup>: $P = 0.055$ vs Stable group (validation cohort).
In the original cohort, the numbers of missing data regarding “history of cigarette smoking”, “temperature $\geq 37^{\circ}\text{C}$ ”, “temperature $\geq 38^{\circ}\text{C}$ ”, “diabetes or obesity”, “hypertension”, “chronic respiratory disease”, “malignancies”, “dyslipidemia”, “cardiac diseases”, and “SpO<sub>2</sub>” were 230, 73, 38, 108, 108, 99, 99, 99, 99, and six, respectively.
In the external validation cohort, the numbers of missing data regarding “diabetes or obesity” was four.
Abbreviations: coronavirus disease 2019, COVID-19; extracorporeal membrane oxygenation, ECMO; mechanical ventilation, MV.

Table 2 shows the results of the forward stepwise multivariate logistic regression analysis that identified independent risk factors associated with worsening COVID-19 during hospitalization. All variables that had significant differences between the groups (Table 1) were included (Model 1, Table 2). Body temperature ≥ 38°C, comorbidities of diabetes or obesity, age ≥ 40 years, and SpO2 < 96% were independent risk factors that predict exacerbation of COVID-19. According to the value of the β regression coefficients of the analysis, the above mentioned variables were assigned points in Model 1 (2 points, 1 point, 1 point, and 1 point, respectively). A model excluding the variable of oxygen saturation was also created (Model 2, Table 3). Body temperature ≥ 38°C, comorbidities of diabetes or obesity, and age ≥ 40 years were independent risk factors that predict the exacerbation (Table 3), and were each assigned 1 point. Both models were significantly associated with the worsening of COVID-19 in the logistic regression analyses (Model 1: *P* < 0.0001, Model 2: *P* < 0.0001). The Hosmer-Lemeshow goodness-of-fit test revealed adequate performances of the predictive models (Model 1: χ^2^ = 1.250, *P* = 0.535; Model 2: χ^2^ = 0.782, *P* = 0.377). The scores in Models 1 and 2 had respective AUCs of 0.789 and 0.771 for predicting the worsening of COVID-19 patients during hospitalization (Figure 2). The optimal cut-off value in both models was 2. The sensitivity and specificity of the cut-off in Model 1 were 70.5% and 77.5%, respectively, and those in Model 2 were 58.2% and 85.7%, respectively. Tables 4 and 5 show the percentages of the patients who had exacerbation (upper) or required the treatment of mechanical ventilation and/or ECMO (lower) during hospitalization according to the scores of Model 1 (Table 4) and Model 2 (Table 5). The higher the points, the more exacerbated and the greater need for mechanical ventilation/ECMO.

**Table 2.** Forward stepwise multiple logistic regression analysis that predicts worsening among mild/moderate-1 COVID-19 patients using all variables that were significantly different between the Stable and Worsened groups (Model 1)

| Variables | $\beta$ regression coefficient | Odds ratio | 95% confidence interval | *P* |
| --- | --- | --- | --- | --- |
| Temperature $\geq 38^{\circ}\text{C}$ | 0.788 | 5.06 | 3.45–7.42 | 1.78E-6 |
| Diabetes or Obesity | 0.536 | 2.98 | 1.91–4.65 | 0.00027 |
| Age $\geq 40$ years-old | 0.523 | 2.66 | 1.72–4.16 | 0.00022 |
| SpO2 < 96% | 0.386 | 2.26 | 1.35–3.76 | 0.00927 |
| Hypertension | — | — | — | 0.078 |
| Positive History of cigarette smoking | — | — | — | 0.141 |
| Temperature $\geq 37^{\circ}\text{C}$ | — | — | — | 0.185 |
| Male gender | — | — | — | 0.598 |

**Table 3.** Forward stepwise multiple logistic regression analysis that predicts worsening among mild/moderate-1 COVID-19 patients using variables that were significantly different between the Stable and Worsened groups except for “SpO2 < 96%” (Model 2)

| Variables | $\beta$ regression coefficient | Odds ratio | 95% confidence interval | *P* |
| --- | --- | --- | --- | --- |
| Temperature $\geq 38^{\circ}\text{C}$ | 0.817 | 5.38 | 3.68–7.86 | 1.22E-6 |
| Diabetes or Obesity | 0.578 | 3.28 | 2.11–5.07 | 5.82E-5 |
| Age $\geq 40$ years-old | 0.535 | 2.74 | 1.78–4.23 | 0.00012 |
| Hypertension | — | — | — | 0.068 |
| Positive History of cigarette smoking | — | — | — | 0.135 |
| Temperature $\geq 37^{\circ}\text{C}$ | — | — | — | 0.129 |
| Male gender | — | — | — | 0.436 |

**Table 4.** Percentages of mild/moderate-1 COVID-19 patients whose conditions worsened during hospitalization (upper, overall worsening; bottom, worsening requiring mechanical ventilation/ECMO) according to the DOATS score (Model 1)

| Point | 0<br>n = 615 | 1<br>n = 515 | 2<br>n = 209 | 3<br>n = 150 | 4<br>n = 42 | 5<br>n = 13 | *P* |
| --- | --- | --- | --- | --- | --- | --- | --- |
| Worsening during hospitalization, % | 2.11 | 5.44 | 14.83 | 29.33 | 38.10 | 53.85 | < 0.0001 |
| Worsening requiring for mechanical ventilation/ECMO, % | 0 | 0 | 0.48 | 0.67 | 7.14 | 15.38 | < 0.0001 |

DOATS score: the prediction of worsening in mild/moderate COVID-19 patients using four items; having diabetes or obesity, age $\geq 40$ years, high temperature of $\geq 38^{\circ}\text{C}$ , and oxygen saturation $< 96\%$ .
These items were assigned 1 point, 1 point, 2 points, and 1 point, respectively. The summation of these points was defined as the patient's DOATS score.

**Table 5.** Percentages of mild/moderate-1 COVID-19 patients who had worsened conditions during hospitalization (upper, overall worsening; lower, worsening requiring for mechanical ventilation/ECMO) according to the DOAT score (Model 2)

| Point | 0<br>n = 646 | 1<br>n = 619 | 2<br>n = 247 | 3<br>n = 36 | *P* |
| --- | --- | --- | --- | --- | --- |
| Worsening during hospitalization, % | 2.32 | 6.95 | 26.32 | 44.44 | < 0.0001 |
| Worsening requiring for<br>mechanical ventilation/ECMO, % | 0 | 0 | 0.81 | 13.89 | < 0.0001 |

DOAT score: the prediction of worsening in mild/moderate COVID-19 patients using three items; having diabetes or obesity, age $\geq 40$ years, and high temperature of $\geq 38^{\circ}\text{C}$ .
Each item was assigned 1 point each. The summation of these points was defined as the patient's DOAT score.

**Figure 2.**
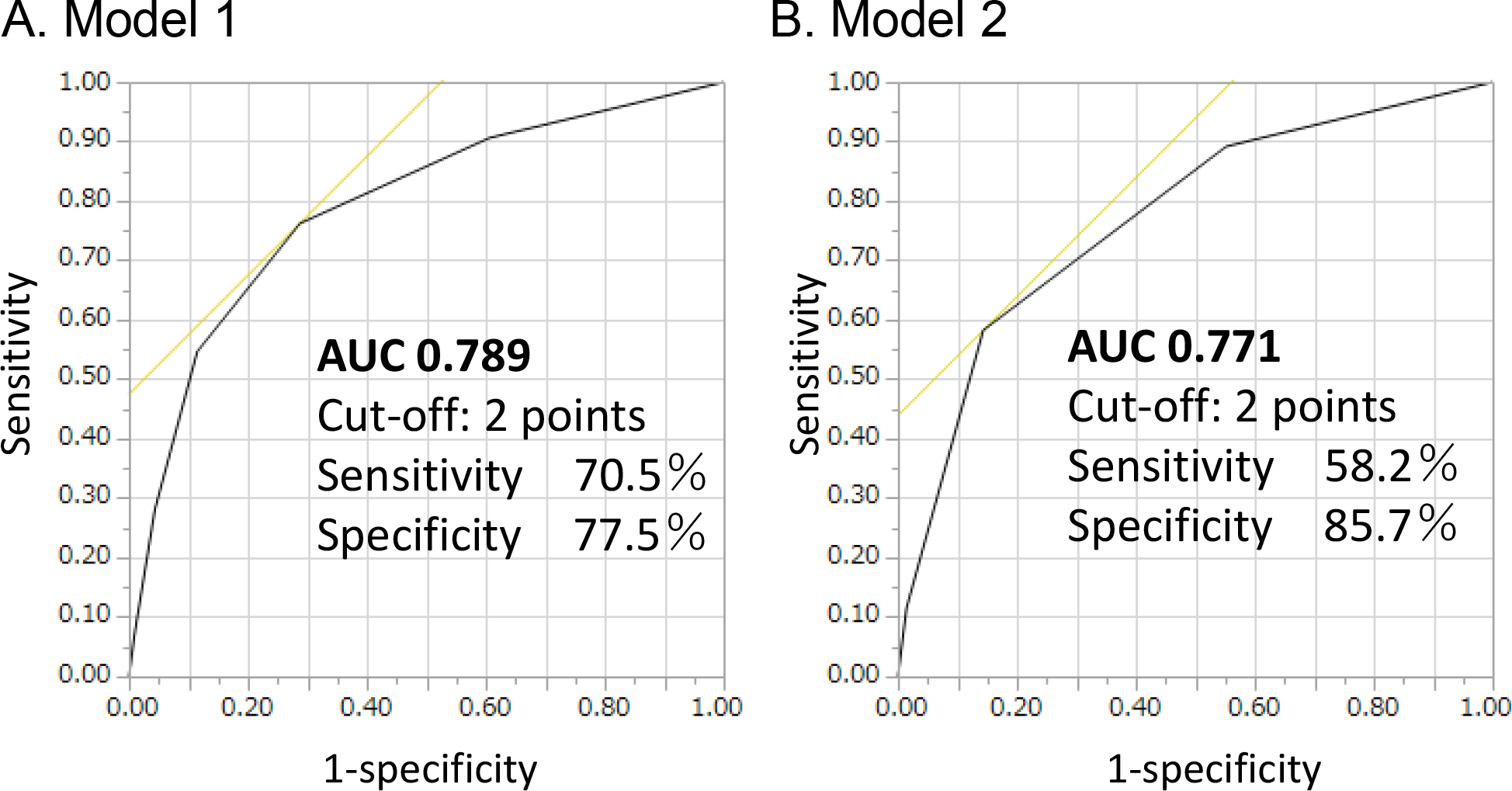
Receiver operating characteristic (ROC) curve analyses of the models that predicts worsening of mild/moderate-1 COVID-19 aged under 65 years. (A) Model 1: prediction model using comorbidity of <u>d</u>iabetes/<u>o</u>bese, <u>a</u>ge ≥ 40 years, body temperature ≥ 38°C, and oxygen saturation < 96% (DOATS score). (B) Model 2: prediction model using comorbidity of <u>d</u>iabetes/<u>o</u>bese, <u>a</u>ge ≥ 40 years, and body temperature ≥ 38°C (DOAT score).

The ability of these models was validated in another cohort. Three hundred and twenty-four inpatients of hospitals in Yamagata, Japan, were enrolled in this study (Table 1). Forty-one of these patients (12.7%) experienced worsening of the disease during hospitalization. In logistic regression analyses, both models were significantly associated with the worsening of COVID-19 (Model 1: *P* < 0.0001, Model 2: *P* < 0.0001). The Hosmer-Lemeshow goodness-of-fit test revealed adequate performances of the predictive models (Model 1: χ^2^ = 5.198, *P* = 0.158; Model 2: χ^2^ = 0.678, *P* = 0.410). The ROC curves of Models 1 and 2 showed respective AUCs of 0.702 and 0.722 for predicting the worsening of COVID-19 with a cut-off of 2, the respective sensitivities were 73.2% and 61.0%, and the respective specificities were 56.6% and 76.7%. Tables 6 and 7 show the percentages of the patients who experienced exacerbation during hospitalization according to the scores of Model 1 (Table 6) and Model 2 (Table 7). The prevalence of worsening outcomes was higher in the high score groups in both models.

**Table 6.** Percentages of mild/moderate-1 COVID-19 patients who had worsening during hospitalization according to the DOATS score (Model 1) in the validation cohort.

| Point | 0<br>n = 79 | 1<br>n = 90 | 2<br>n = 68 | 3<br>n = 42 | 4<br>n = 31 | 5<br>n = 10 | *P* |
| --- | --- | --- | --- | --- | --- | --- | --- |
| Worsening during hospitalization, % | 3.80 | 8.89 | 10.29 | 30.95 | 25.81 | 20.00 | < 0.0001 |

Due to four missing data, this analysis was performed using the data of 320 patients. Data regarding the utilization of a mechanical ventilator or ECMO were not available in the validation cohort.
DOATS score: the prediction of worsening in mild/moderate COVID-19 patients using four items, having of diabetes or obesity, age ≥ 40 years, high temperature $\geq 38^{\circ}\text{C}$ , and oxygen saturation $< 96\%$ .
These items were assigned 1 point, 1 point, 2 points, and 1 point, respectively. The summation of these points was defined as the patient's DOATS score.

**Table 7.**
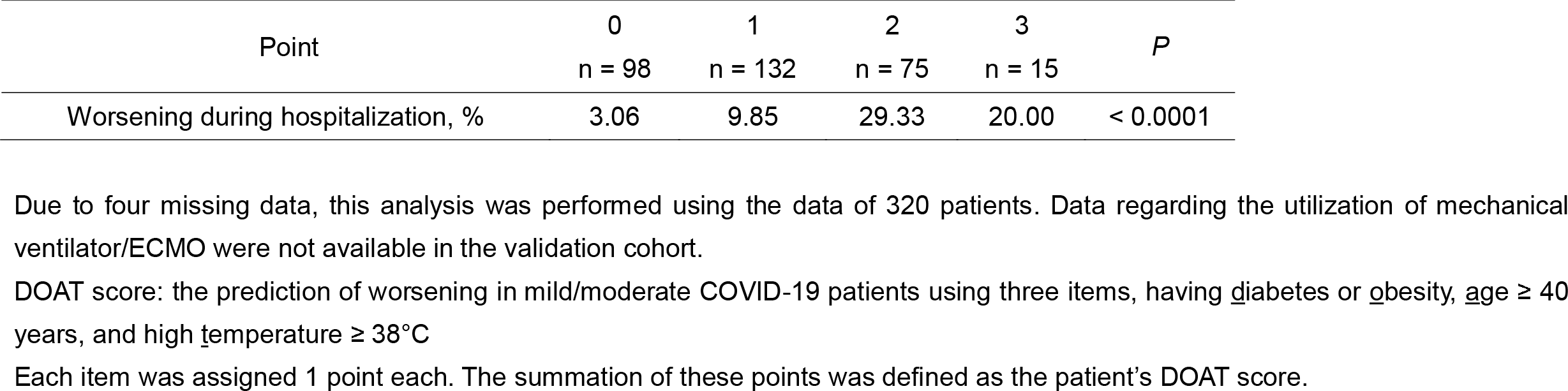
Percentages of mild/moderate-1 COVID-19 patients who had worsening during hospitalization according to the DOAT score (Model 2) in the validation cohort.

## Discussion

We examined risk factors associated with worsening COVID-19 in 1,675 Japanese patients without respiratory failure aged < 65 years. Using the identified risk factors, we established two predictive models; one consisting of four items, namely comorbidity of <u>d</u>iabetes or <u>o</u>besity, <u>a</u>ge ≥ 40 years, high body temperature ≥ 38°C, and oxygen saturation < 96%, DOATS score; and another consisting of three items, <u>d</u>iabetes or <u>o</u>besity, <u>a</u>ge, and temperature, DOAT score. In cases where the doctor visited the patients’ homes, information about oxygen saturation may not be available. Therefore, we thought that prediction using three items, DOAT score, is easier to use than that consist of four items, the DOATS score. Because the difference of the area under ROC curve between the two models was less than 2%, we believe that their predictive abilities were not significantly different.

The present study included a relatively large number of subjects in Fukushima Prefecture. COVID-19 is a designated “Category 2 Infectious Disease” in Japan, and in Fukushima Prefecture, inpatient treatment was performed for even mild cases, in order to isolate the patients from uninfected persons. Many COVID-19 studies have been primarily from the in-patient setting, skewing toward severe disease. In our database, not only severely ill patients but also patients with mild and moderate cases, who did not have respiratory failure, were enrolled. As a result, over 1,500 patients who did not have respiratory failure were included in the analysis.

In the present study, clinical worsening in each patient was identified based on data regarding starting medication against COVID-19, oxygen inhalation including nasal high flow cannula, and mechanical ventilation. This information enabled us to understand the risk factors that predict worsening COVID-19 among patients in the early phase of the disease. Most previous studies have analyzed either risk factors that predict death or mechanical ventilator used mainly in critically ill COVID-19 patients[2-7]. However, it is unclear whether the predictive factors identified in those studies can be used to accurately identify patients with mild cases who may experience worsening, because the statistical analyses of those studies only focused on the extremely hard outcomes such as death or use of mechanical ventilator. Even if the disease progression does not lead to such a hard outcome, the progression to respiratory failure often has a significant effect on the patient. Some patients with respiratory failure may require hospitalization for a long period of time, and may require long-term oxygen therapy due to the lowering of pulmonary function. Therefore, prevention of progression to respiratory failure is considered to be a major medical goal. For this purpose, it is necessary to identify patients who are at risk of exacerbation and provide treatment such as neutralizing antibodies or antiviral agents to those individuals as early as possible[22, 23].

Several studies demonstrated the association between the clinical characteristics such as older age, male sex, symptoms of COVID-19 (fever, cough, fatigue, and short of breath) and comorbidities (hypertension, diabetes, and CKD) and its severity or activity[24-26]. Addition of blood test may increase the accuracy of prediction of the disease progression[10]. In the present study, we demonstrated two very simple methods that predict the development of illness among mild/moderate patients with COVID-19, DOAT and DOATS scores. Because blood sampling is not needed for these scorings, any medical staffs can evaluate the risk of patient in front of them immediately. Tu et al. made a model predicting progression of COVID-19 consisting of nine items, namely male sex, age, fever, hypertension, cardio-cerebrovascular disease, dyspnea, cough, and myalgia [13]. Their model consisted of clinical characteristics only, not laboratory data. Their ability to predict the progression to the disease seemed to be equal to ours because the areas under the ROC curve was 0.787 in their model, and 0.790 (DOATS score) and 0.771 (DOAT score) in our models. However, the models demonstrated in the present study was simpler than their model. During the situation of the pandemic where many patients are coming to clinics, the simple and quick methods to identify the worsening patients is desirable.

Currently, many elderly people in Japan have been already vaccinated against SARS-CoV-2 [27, 28]. Delayed vaccination for middle aged and younger people due to a vaccine shortage resulted in the shift of epidemics from elderlies to middle-aged/young people. We suspect that this change means that there are different risk factors of worsening COVID-19 to identify and assess. Previously reported risk factors may not be effective for the groups of people who are currently at highest risk, because comorbidities such as cancer and chronic respiratory or renal diseases are relatively uncommon in young and middle-aged people. In the present study, the risk factors for worsening COVID-19 among non-elderly patients without respiratory failure were comorbidities of diabetes or obesity, high body temperature, older age, and lower oxygen saturation. The reported risks, such as male sex, cigarette smoking, hypertension, chronic respiratory disease, malignancies, dyslipidemia, and cardiac diseases, were not independently associated with worsening of the disease. Therefore, the risk factors for worsening COVID-19 may need to be reconsidered and repeatedly updated in accordance with the shift of generations who are suffering from COVID-19.

The strength of the current study is that the results are considered to be generalizable, because this study included inpatients at major hospitals in Fukushima that can handle COVID-19 inpatient treatment. In our database, three fourths of COVID-19 patents in Fukushima were enrolled. In addition, the robustness of our results is supported by the results from the external validation cohort analyses.

There were limitations in the present study. Firstly, data regarding the exact proportion of vaccinated individuals were not available. COVID-19 vaccinations began to be administered in Japan in March 2021, and by the end of May 2021, no vaccinations had yet been administered to non-elderly people [27, 28]. Therefore, we believe that most of the population in this study had not been vaccinated. Secondly, the precise data regarding about the day of onset and deterioration or treatment before deterioration was not available for any patients.

There may be some differences in treatment before deterioration between the Worsened and Stable groups. For example, treatment such as inhaled corticosteroid and favipiravir may have affected the clinical course of the patients[29-31]. In our database, information about the timing of the prescription of these medicines were not available. Thirdly, we do not know whether the risk factors identified in the present study will still be applicable to the risk stratification against COVID-19 caused by future-coming variants of SARS-CoV-2. The current vaccines are reportedly less effective against the SARS-CoV-2 Mu variant[32]. Therefore, even vaccinated individuals may still get infected[33]. The increased infectivity caused by viral mutations may prolong the COVID-19 pandemic. In situations where many individuals are vaccinated, it is possible that new risk factors for worsening COVID-19 will become apparent.

## Conclusion

We found that comorbidities of diabetes or obesity, age ≥ 40 years, high body temperature ≥ 38°C, and oxygen saturation < 96% were risk factors which were linked to disease progression in non-elderly COVID-19 patients who did not have respiratory failure on admission. We also established two simple prediction models that can quickly and easily evaluate the risk of the patients using the sum of the points that were given according to the presence of these risk factors. We believe that these scoring methods can be used broadly in many clinics, and that they can be used to effectively identify high risk patients among non-elderly COVID-19 patients at an early phase, resulting in reducing progression of the disease by enabling treatment to be administered as soon as possible.

## Data Availability

All data produced in the present work are contained in the manuscript.

## Footnote page

### Funding

This study received no specific grant from any funding agency.

#### Abbreviations

COVID-19: coronavirus disease 2019
DOAT: comorbidity of <u>d</u>iabetes/<u>o</u>besity, <u>a</u>ge ≥ 40 years, and body temperature ≥ 38°C
DOATS: comorbidity of <u>d</u>iabetes/<u>o</u>besity, <u>a</u>ge ≥ 40 years, body temperature ≥ 38°C, and oxygen saturation < 96%
ECMO: extracorporeal membrane oxygenation
SARS-CoV-2: severe acute respiratory syndrome coronavirus 2

## Acknowledgements

We thank the Scientific English Editing Section of Fukushima Medical University for their fruitful discussion and linguistic assistance in proofreading the manuscript.

