## Supplementary Methods for "Development and external validation of the DOAT and DOATS scores: simple decision support tools to identify disease progression among nonelderly patients with mild/moderate COVID-19"

^1^Department of Pulmonary Medicine, Fukushima Medical University; ^2^Department of Pulmonary Medicine, Ohara General Hospital; ^3^Department of Thoracic Surgery, Fukushima Red Cross Hospital; ^4^Department of Internal Medicine, Fujita General Hospital; ^5^Department of Pulmonary Medicine, Saiseikai Fukushima General Hospital; ^6^Department of General Internal Medicine and Clinical Infectious Diseases, Fukushima Medical University; ^7^Department of Internal Medicine, Saiseikai Kawamata Hospital; ^8^Department of Emergency and Critical Care Medicine, Aizu Chuo Hospital; ^9^Department of Infectious Disease and Pulmonary Medicine, Aizu Medical Center, Fukushima Medical University; ^10^Department of Internal Medicine, Takeda General Hospital; ^11^Department of Pediatric Medicine, Bange Kousei General Hospital; ^12^Department of Surgery, Yurin Hospital; ^13^Department of Pediatric Surgery, Iwaki City Medical Center; ^14^Department of Internal Medicine, Kashima Hospital; ^15^Department of Neurosurgery, Fukushima Rosai Hospital; ^16^Department of Emergency and Critical Care Medicine, Futaba Medical Center; ^17^Department of Internal Medicine, Soma General Hospital; ^18^Department of Pulmonary Medicine, Minami-Soma Municipal General Hospital; ^19^Department of Surgery, Onahama Chuo Clinic; ^20^Department of General Medicine, Shirakawa Satellite for Teaching and Research, Fukushima Medical University; ^21^Department of Cardiology and Vascular Medicine, Hoshi General Hospital; ^22^Department of Internal Medicine, Iwase General Hospital; ^23^Department of Surgery, Southern TOHOKU General Hospital; ^24^Department of Pulmonary Medicine, Jusendo General Hospital; ^25^Department of Emergency and Critical Care Medicine, Ohta Nishinouchi Hospital; ^26^Department of Respiratory Medicine, Tsuboi Hospital; ^27^Department of Pulmonary Medicine, Okitama General Hospital; ^28^Department of Pulmonary Medicine, Nihonkai General Hospital; ^29^Department of Pulmonary Medicine, Yamagata City Hospital Saiseikan; ^30^Department of Emergency and Critical Care Medicine, Fukushima Medical University; ^31^Department of Innovative Research and Education for Clinicians and Trainees, Fukushima Medical University Hospital

**Corresponding author**

Yoko Shibata

Department of Pulmonary Medicine, Fukushima Medical University School of Medicine, Fukushima, 960-1295, Japan.

**Supplementary Methods**

Statistical analyses

The variables that have statistical significant differences between the two groups were used to identify independent risk factors for predicting the worsening of COVID-19 a day or later after admission to hospital by forward stepwise multivariate logistic regression analyses. Because information on oxygen saturation may not be available in some clinical sites such as the patients’ temporary accommodation or homes, we analyzed two models; one including and one excluding oxygen saturation values. Points proportional to the beta regression coefficient values (rounded to the nearest integer) were assigned to each variable to derive simple clinical prediction scores, and the effectiveness of the prediction models were confirmed by the area under receiver operating characteristic (ROC) curve values. The Hosmer-Lemeshow goodness-of-fit test was used to evaluate the agreement between observed and predicted outcomes with respect to calibration ability of the scores[1]. Sample size calculations were not performed because we used all available data on the registry to maximize the power of the results.

For external validation, patients with COVID-19 treated in three hospitals in Yamagata Prefecture were used. Information on age, gender, presence of diabetes and/or obesity, body temperature, and oxygen saturation levels were provided from nonelderly patients with mild/moderate-1 COVID-19. Associations between the outcomes and the two derived clinical prediction scores were examined using logistic regression analyses, and area under ROC curve values were calculated. All statistical analyses were performed using JMP 13 (SAS Institute Inc, Cary NC) and SPSS27 (IBM Corp., Armonk, NY). A two-tailed p-value of < 0.05 was considered statistically significant. Cases with missing data were not excluded from analyses, and the details of the missing data are described in the note of Table 1.
